## Appendix A: Interview Guide for "Development of a core outcome set for multimorbidity trials in low- and middle-income countries (COSMOS): Study Protocol"

### Development of a core outcome set for multimorbidity studies in low- and middle-income countries (COSMOS)

### Interview Guide for Patients

**Version: 1.3**
**Date: 28-10-20**

*Demographics (age, gender, marital status, highest level of education, socioeconomic status, occupation and disability should be recorded at eligibility checking/recruitment) need to be collected on a separate 'demographics' sheet.*

***Introduction:***

Our goal is to understand what outcomes are important to you in preventing or treating co-existing chronic illnesses. We also want to identify what terms you might prefer to use to describe these outcomes.

What is an outcome?

To help patients, their family, doctors and other health professionals make decisions about treatments, we need evidence about what works best. Treatments are developed and tested by researchers to make sure they work and are safe. To do this, researchers need to look at the effects those treatments have on patients.

Researchers do this by measuring an 'outcome'. For example, in a study of how well a new asthma treatment works, 'outcomes' might include: a measure of how fast the asthma attack gets better; how many times a person has to stay in hospital because of asthma; or how well a person can get on with their usual activities because of asthma.

What is a core outcome set?

If all studies of a particular health condition(s) use the same outcomes, they can all be compared and combined. When a set of main outcomes has been agreed for a health condition(s), it's called a 'core outcome set'. If studies use these core outcomes, we could bring together all the studies to get a better understanding of which treatments are best.

What do we need you to tell us?

Core outcomes have to be relevant to patients, family members as well as health professionals and researchers. Some outcomes may be more important to patients and families than to healthcare professionals but without asking we will not know this and they might not get included in the core outcome list.

[Interviewer, before proceeding to the interview please check (if not already done):
- whether the participant information sheet had been shared and discussed;
- whether consent was obtained and documented.]

*Interview:*

*As a quick reminder, we are interested in your experience of living with [living with someone living with] two or more health conditions at the same time. This is what we mean when using the terms "co-existing" or "multimorbidity". With health conditions we mean mental disorders, non-communicable diseases, and/or communicable diseases. In this interview we are interested in your view on what matters to you as the result of treatment and/or care for your conditions.*

1. Please can you tell me how long (approximately) you [your family member] have had these co-existing chronic illnesses?

2. Where have you received treatment for these?
*Probes: In the community, primary care, hospital, tertiary centre.*

3. (For family members only) What is your relationship to your family member with these conditions?

4. Please tell me a little bit about your (your family member's) experience living with these conditions
*Probe: What matters to you the most about the impact of these conditions?*

5. What do you (your family member) hope to gain through treatment or care?

6. Speaking from your experience, how do you know that a treatment for your conditions is working?

*[Interviewer, to finish off this question, ask]*: What would you say your priorities are?

7. Considering this experience, what would you want future studies of prevention and treatment to look at in order to make it better for others in the future?

8. Is there anything else you'd like to talk about in relation to research for treatments to prevent or treat these conditions?
